# Association between dietary inflammatory index score and incident dementia: results from the Framingham heart Study offspring cohort

**DOI:** 10.1101/2023.08.21.23294374

**Authors:** Debora Melo van Lent, Hannah Gokingco Mesa, Meghan I. Short, Mitzi M. Gonzales, Hugo J Aparicio, Joel Salinas, Changzheng Yuan, Paul F. Jacques, Alexa Beiser, Sudha Seshadri, Mini E. Jacob, Jayandra J. Himali

## Abstract

**Background:** The Dietary Inflammatory Index (DII), has been specifically designed to capture the inflammatory content of diet and has shown association with neurodegenerative disease related outcomes. But literature is limited on the role of diet-driven inflammation measured by the DII on incident all-cause dementia and Alzheimer’s disease dementia (AD).

**Objective:** We evaluated whether higher DII scores were associated with increased incidence of all-cause dementia and AD over 22.3 years of follow-up in the community-based Framingham Heart Study (FHS) Offspring cohort.

**Design, Setting, and Participants:** Observational longitudinal study in the FHS Offspring cohort. Dementia surveillance for present study: until 2020. Data were analyzed from December 2020 to June 2022. Participants completed a validated 126-item food frequency questionnaires (FFQ), administered at FHS examination cycle 7 (1998-2001) and examination cycle 5 (1991-1995), and/or 6 (1995-1998). Individuals aged <60 years, with prevalent dementia, no dementia follow-up, other relevant neurological diseases, and/or no FFQ data were excluded.

**Exposure:** A DII score (based on the published method by Shivappa et al. 2014) was created based on previous studies linking individual dietary factors to six inflammatory markers (i.e. C-reactive protein, interleukin (IL)-1β, IL-4, IL-6, IL-10, and tumor necrosis factor-alpha), consisting of 36 components. A cumulative DII score was calculated by averaging across a maximum of three FFQs.

**Main outcomes and measures:** Incident all-cause dementia and AD.

**Results:** We included 1487 participants (mean±SD, age in years 69 ± 6; 53·2% women; 31·6% college graduates]). 246 participants developed all-cause dementia (including AD n=187) over a median follow up time of 13·1 years. Higher DII scores were associated with an increased incidence of all-cause dementia and AD following adjustment for age and sex (Hazard ratio (HR) 1·16, 95% confidence interval (CI) 1·07 to 1·25, p<.001; HR 1·16, 95% CI 1·06 to 1·26, p=.001). The relationships remained after additional adjustment for demographic, lifestyle, and clinical covariates (HR 1·21, 95% CI 1·10 to 1·33, p<0.001; HR1·20, 95% CI1·07 to 1·35, p=.001).

**Conclusion and relevance:** Higher DII scores were associated with a higher risk of incident all-cause dementia and AD. Although these promising findings need to be replicated and further validated, our results suggest that diets which correlate with low DII scores may prevent late-life dementia.

## Introduction

Dementia is currently one of the biggest global health and social care challenge with an estimated 152 million persons predicted to be living with dementia worldwide by 2050.^1^ To address this unprecedented burden of disease, the global focus has been on developing multidisciplinary and personalized care for individuals with dementia and escalating research funding for dementia drug trials.^2,3^ Although this approach has led to better care for individual patients, the licensing of five drugs that treat dementia symptoms, and two drugs which target the underlying biology of Alzheimer’s disease ^3^, there has been limited progress in identifying effective interventions that actually prevent, stop or reverse the progression of dementia. At this juncture, there is a pressing need to re-consider primary prevention strategies, starting with the identification of modifiable risk factors, such as diet.^4^

Dietary interventions have shown to be successful in combating chronic disease related risk factors. For example, a meta-analyses of nutrition education and Mediterranean (style) diet randomized control trials (RCT) targeting type 2 diabetes, have reported positive effects on clinical parameters of glycemic control, body weight and cardiovascular risk factors.^5,6^ Dietary interventions are now widely employed in clinical settings and as part of broader population level public health strategies to prevent and reduce chronic disease burden. Successful outcomes from these trials have encouraged investigation into the potential role of dietary intervention in dementia prevention as well. For instance the Prevención con Dieta Mediterránea (PREDIMED) and PREDIMED-Plus RCTs have shown that the Mediterranean diet can improve cognition.^7,8^ Additionally, a Mediterranean-Dietary Approach to Systolic Hypertension (DASH) diet intervention for neurodegenerative delay (MIND) diet RCT found a beneficial effect of the diet on cognitive performance and brain structure.^9^ These trials indicate the potential scope of dietary interventions for prevention of dementia.

Beneficial effects of dietary intake intervention studies on cardiometabolic and cognitive health have been attributed to their pivotal role in the regulation of systemic inflammation, one of the hypothesized contributary pathways leading to dementia.^10,11^ We have evidence that the Mediterranean diet significantly lowered concentrations of inflammatory markers in the PREDIMED RCT.^12^ These circulating inflammatory markers, have, in turn, been associated with incident all-cause dementia and probable AD dementia in population-based studies.^13,14^ The investigation of the direct relationship between the inflammatory content of diet and risk of dementia is key to understanding the role of diet on the inflammatory pathway to dementia. If causal relationships are found between the inflammatory potential of diet and neurodegenerative diseases, these findings will have implications for the optimization of existing dietary guidelines.

The Dietary Inflammatory Index (DII), is an index that has been specifically designed to capture the inflammatory potential of diet ^15^ and has shown association with neurodegenerative disease related outcomes in previous studies.^16–22^ However there has been limited information on the longitudinal relationship between diet-driven inflammation measured by the DII and incident all-cause and Alzheimer’s disease (AD) dementia.^16,23^ One previous study was limited to women and the second one had a maximum follow-up time of 6.08 years and examined all-cause dementia incidence only. ^23^ A large cohort including men and women, with longer follow-up time and usual dietary intake measured over consecutive visits would provide substantial evidence for the role of diet in changing the risk for all-cause and AD dementia.

In order to evaluate previously identified links between dietary inflammatory content and risk of dementia, we examined the relationship in a large established cohort with repeated diet measures and extensive follow-up time. We tested the association between the DII and incident all-cause and AD dementia over a period up to 22.3 years follow-up in 1487 participants of the community-based FHS Offspring cohort. A positive association if detected would imply that dietary intervention could be part of public health preventive strategies for dementia in large populations.

## Methods

The FHS is an ongoing community-based study of several cohorts from the town of Framingham, Massachusetts, USA (https://www.framinghamheartstudy.org/). The Original cohort was established in 1948 with the aim to identify factors that contribute to the development of cardiovascular disease.^24^ In 1971, the Offspring cohort was established, including children of the Original cohort and their spouses.^25^ The Offspring cohort enrolled 5124 participants who have been studied over nine examination cycles, approximately once every 4 years. All participants provided written informed consent. The study protocol was approved by the institutional review board at Boston University Medical Center.

For the present study, we assessed self-reported dietary intake from participants of the Offspring cohort using a food frequency questionnaire (FFQ) administered at examination cycles 5 (1991-1995), 6 (1995-1998), and 7 (1998– 2001). A flow chart of sample selection is shown in Figure 1. To be included in the present investigation, participants were required to have completed the FFQ at examination cycle 7 and at least one other time point (examination cycles 5 or 6). We commenced dementia follow-up from examination cycle 7. Participants were excluded if they had no dietary intake data available or an abnormal estimated total energy intake (<600 - >3999 kcal for women or <600 - >4199 kcal for men), and/or >13 missing FFQ items (n=643).^26^ We excluded participants aged <60 years (n=1326), with prevalent dementia (n=10), and with no dementia follow-up (n=73).

**Figure 1.**
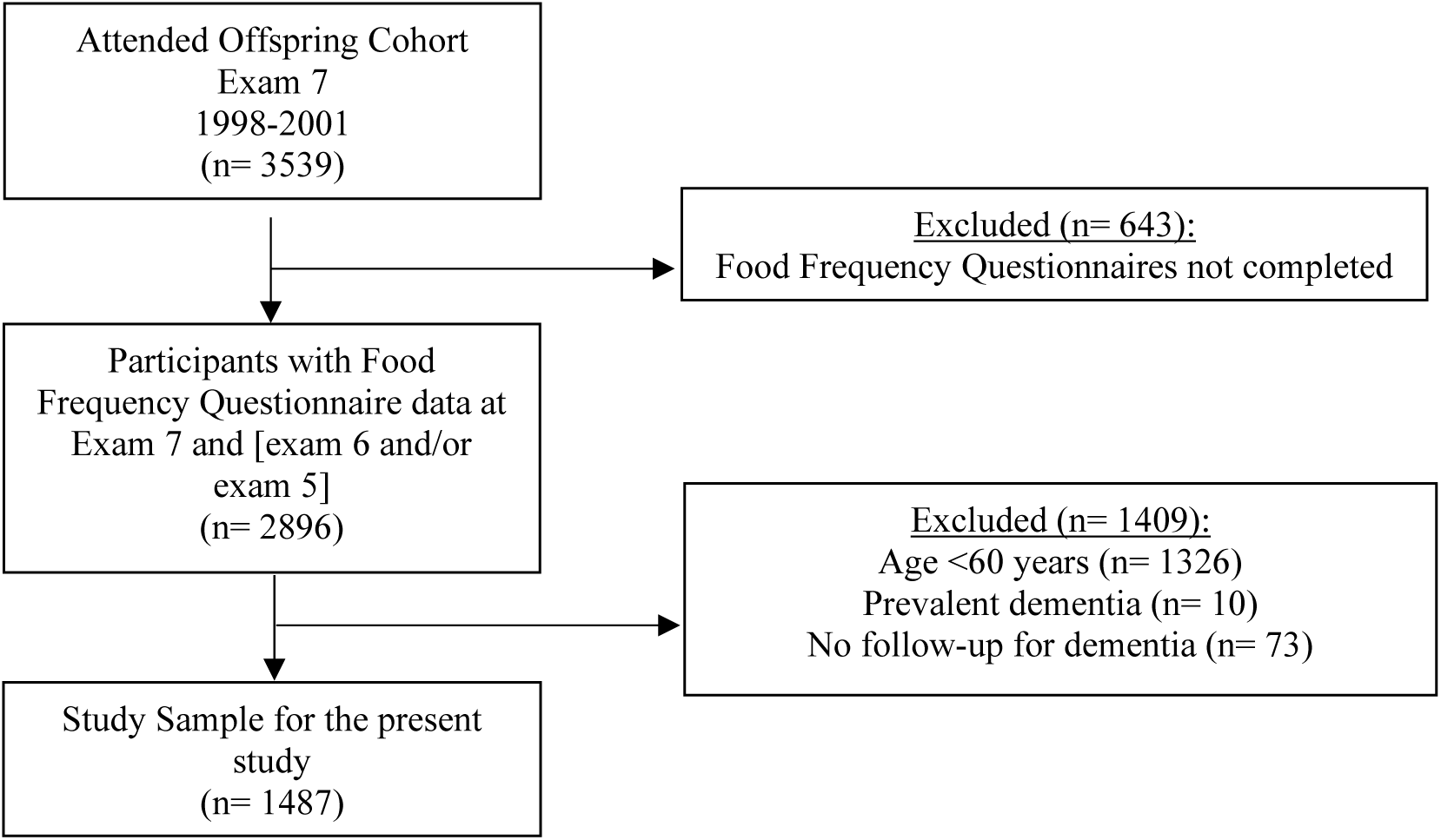
Flow chart of the participants included in the study.

### Dietary inflammatory index scores

The DII index for the present study was calculated using the validated 131-item Harvard semi-quantitative FFQ.^27,28^ The FFQ assesses dietary intake over the past year. Participants were asked how often they consumed food items (i.e., from never or <1 per month to >6 per day), the type of food item and whether food items were homemade or ready-made ^29^. A commonly used portion size was given for each food item. In addition, the FFQ included questions on most frequently eaten breakfast cereal, types of fats and oils, and frequency of consumption of fried foods. Intakes of food components (i.e. nutrients, food items or food groups) were computed by multiplying the frequency of consumption of each food item by the nutrient content of the specified portions.^29^

The DII index for the present study consists of 36 dietary components including anti-inflammatory nutrients, pro-inflammatory nutrients, whole foods and caffeine (5). The dietary components are categorized as (1) *Anti-inflammatory*: alcohol, beta-carotene, caffeine, dietary fiber, folic acid, magnesium, thiamin, riboflavin, niacin, zinc, monounsaturated fat, polyunsaturated fat, omega-3 fat, omega-6 fat, selenium, vitamins B6, A, C, D, E, flavan-3-ol, flavones, flavonols, flavonones, anthocyanidins, green/black tea, pepper, and garlic; and (2) *pro-inflammatory*: vitamins B12, iron, carbohydrates, cholesterol, total energy intake, protein, saturated fat, and total fat. Additionally, total energy intake was categorized as pro-inflammatory. Nine components of the Shivappa et al. 2014 DII (i.e. turmeric, thyme/oregano, rosemary, eugenol, ginger, onion, transfat, isoflavones, and saffron) were not available in our FFQ. The algorithm for the DII computation has been described elsewhere **(eTable 2 in the supplement)**.^15^

At FHS we had the opportunity to assess dietary intake over a decade. As data from a FFQ is prone to recall bias, and to account for reverse causality, we averaged the DII scores across examination cycle 7 (1998-2001) and at least one of the prior examination cycles: 5 (1991-1995) and 6 (1995-1998).

### Ascertainment of incident all-cause dementia and AD dementia

Incident all-cause dementia and AD dementia were ascertained through continuous surveillance in FHS^30^. In short, the Offspring cohort participants were screened at each examination visit via the MMSE since 1991. Participants with suspected cognitive impairment who do not meet diagnostic criteria for dementia undergo regular neuropsychological assessments between the scheduled examinations. Participants flagged for possible cognitive impairment or dementia on this testing are subsequently evaluated by a neurologist who may refer the participants file for administrative dementia review. A diagnosis of dementia is based on a review of all available neurologic examination records, neuropsychological assessments, neuroimaging investigations, hospital/nursing home/outpatient clinic records, family interviews, and autopsy results (when available) by a dementia review committee that includes a minimum of one neuropsychologist and a neurologist. If a participant dies or does not attend further follow-up, the dementia review committee reviews medical records up to the time of death/loss to follow-up, to ascertain if the participant may have developed interval cognitive impairment/dementia since the last study examination. All-cause dementia was diagnosed using criteria from the Diagnostic and Statistical Manual of Mental Disorders, 4th Edition (DSM-IV) ^31^ and AD dementia was diagnosed based on the criteria of the National Institute of Neurological and Communicative Disorders and Stroke and the AD and Related Disorders Association (NINCDS-ADRDA) for possible or probable AD.^32^

### Statistical analysis

SAS Software 9.4 (SAS Institute, Cary, NC, USA) was used to perform a series of Cox proportional hazard models to examine the longitudinal relationships between DII scores and incident all-cause and AD dementia. The proportional hazards assumption was upheld. We confirmed the proportionality of hazards by including time dependent covariates in the Cox model. Time dependent covariates were created as interactions of the predictors and function of survival time and included in the model. Main results are presented as adjusted hazard ratios accompanied by 95% confidence intervals. The hazard ratios represent the change in all-cause and AD dementia risk by each unit increase in the DII scores. A *P*-value <0.05 and <0.10 was considered statistically significant for the main analyses and for the tests of interactions, respectively. Missing data were excluded from analysis. Confounders were selected based on the published literature **(eTable 3 in the Supplement)**. Model 1 was adjusted for age, and sex. Model 2 was additionally adjusted for education, Apolipoprotein ε4 status (*APOE* ε4), body mass index (BMI), smoking status, physical activity index score, total energy intake, use of lipid-lowering medication, and total cholesterol to high-density lipoprotein (HDL) cholesterol ratio. For our secondary analyses, we tested for interactions between the DII score and *APOE* ε4 status, sex, hypertension, prevalent diabetes mellitus type 2, and cardiovascular disease separately using model 2.

## Results

### Cohort demographics

**Table 1** details the sample characteristics. The mean DII score was -0·30 ± 1·69, indicating that this sample’s diets were on average anti-inflammatory relative to the global mean. Over a mean follow-up time of 12·8 ± 6 years, up to 22·3 years; 246 participants developed all-cause dementia, of which 187 individuals developed AD dementia.

**Table 1.** Baseline characteristics of participants included in the study.

|  | Overall | DII Q1 | DII Q2 | DII Q3 | DII Q4 |
| --- | --- | --- | --- | --- | --- |
| Variable | (n=1487) | (n=371) | (n=372) | (n= 372) | (n= 372) |
| Age, years | 69 ± 6 | 69 ± 6 | 69 ± 6 | 68 ± 5 | 68 ± 6 |
| Follow-up time in years | 12.8 ± 6.0 | 13.1 ± 6.0 | 12.8 ± 6.0 | 12.8 ± 6.1 | 12.3 ± 6.1 |
| Follow-up time in years, median (IQR) | 13.1 (7.9-18.2) | 13.5 (8.1-18.5) | 13.4 (8.5-17.9) | 12.9 (7.7-18.4) | 12.8 (7.4-18.0) |
| Men, n (%) | 696 (46.8) | 163 (43.9) | 180 (48.4) | 174 (46.8) | 179 (48.1) |
| Apolipoprotein ε4 allele, n (%) | 328 (22.3) | 76 (20.6) | 84 (23.0) | 90 (24.5) | 78 (21.3) |
| Education, n (%) |  |  |  |  |  |
| No high school degree | 87 (5.9) | 15 (4.1) | 24 (6.5) | 21 (5.8) | 27 (7.3) |
| High school degree | 492 (33.6) | 90 (24.7) | 106 (28.8) | 131 (35.9) | 165 (44.7) |
| Some college | 424 (28.9) | 108 (29.7) | 108 (29.4) | 110 (30.1) | 98 (26.6) |
| College graduate | 463 (31.6) | 151 (41.5) | 130 (35.3) | 103 (28.2) | 79 (21.4) |
| BMI, kg/m<sup>2</sup> | 28.0 ± 4.9 | 27.4 ± 4.7 | 27.8 ± 4.6 | 28.6 ± 5.2 | 28.3 ± 4.9 |
| PA Index | 38.1 ± 6.4 | 38.4 ± 6.1 | 38.0 ± 6.4 | 38.4 ± 6.2 | 37.9 ± 6.7 |
| Total energy intake, kcal | 1796 ± 557 | 2175 ± 518 | 1869 ± 484 | 1716 ± 499 | 1424 ± 446 |
| Current smoker, n (%) | 115 (7.7) | 11 (3.0) | 23 (6.2) | 30 (8.1) | 51 (13.7) |
| Total cholesterol, mg/dL | 199 ± 36 | 197 ± 36 | 197 ± 36 | 201 ± 35 | 201 ± 38 |
| HDL cholesterol, mg/dL | 53 ± 17 | 54 ± 18 | 54 ± 17 | 53 ± 17 | 52 ± 17 |
| Total to HDL Cholesterol ratio | 4.0 ± 1.2 | 4.0 ± 1.3 | 3.9 ± 1.1 | 4.1 ± 1.3 | 4.1 ± 1.2 |
| Lipid-lowering medication, n (%) | 395 (26.6) | 89 (24.0) | 93 (25.0) | 112 (30.2) | 101 (27.2) |
| Prevalent Diabetes Mellitus type 2, n (%) | 232 (15.7) | 52 (14.1) | 67 (18.0) | 59 (15.9) | 54 (15.6) |
| Prevalent CVD, n (%) | 273 (18.4) | 75 (20.2) | 70 (18.8) | 58 (15.6) | 70 (18.8) |
| JNC VII Stage I Hypertension, n (%) | 861 (57.9) | 213 (57.4) | 201 (54.0) | 224 (60.4) | 223 (60.0) |
| DII score | -0.30 ± 1.69 | -2.48 ± 0.65 | -0.91 ± 0.35 | 0.32 ± 0.38 | 1.88 ± 0.68 |
| All cause dementia cases, n (%) | 246 (16.5) | 50 (13.5) | 60 (16.1) | 69 (18.6) | 67 (18.0) |
| Alzheimer's disease dementia cases, n (%) | 187 (12.6) | 38 (10.2) | 47 (12.6) | 50 (13.4) | 52 (14.0) |

Mean ± SD reported, unless specified otherwise.
Abbreviations: BMI= body mass index; CVD= cardiovascular disease; DII= dietary inflammatory index; dL= deciliter; HDL= high-density lipoprotein; IQR= interquartile range; JNC VII Stage I= The Seventh Report of the Joint National Committee on Prevention, Detection, Evaluation, and Treatment of High Blood Pressure Stage I; kcal= kilocalorie; kg= kilogram; L=liter; m=meter; mmol=millimole; mg= milligram; mL=milliliter; n= number of participants; ng= nanograms; PA= physical activity; Q= quartile.

### Dietary inflammatory index scores and incident all-cause and AD dementia

Higher (pro-inflammatory) DII scores were linearly associated with an increased incidence of all-cause dementia after adjustment for age and sex (Model 1: hazard ratio (HR) = 1·16, 95% CI 1·07 ; 1.25, p< .001) **(Table 2)**. After additional adjustment for education, *APOE* ε4, total energy intake, BMI, smoking status, physical activity index score, total cholesterol to HDL cholesterol ratio, and the use of anti-cholesterol medication the association remained (Model 2: 1·21 (1·10; 1·33), p< .001). In addition, in comparison to the first quartile (i.e. most anti-inflammatory), the third and fourth DII score quartiles (i.e. most pro-inflammatory) were associated with increased incidence of all-cause dementia (Model 2: quartile 3: 1·67 (1·10; 2·52), p =.02; quartile 4: 1·84 (1·18; 2·89), p= .01; p for trend= .04).

**Table 2.**
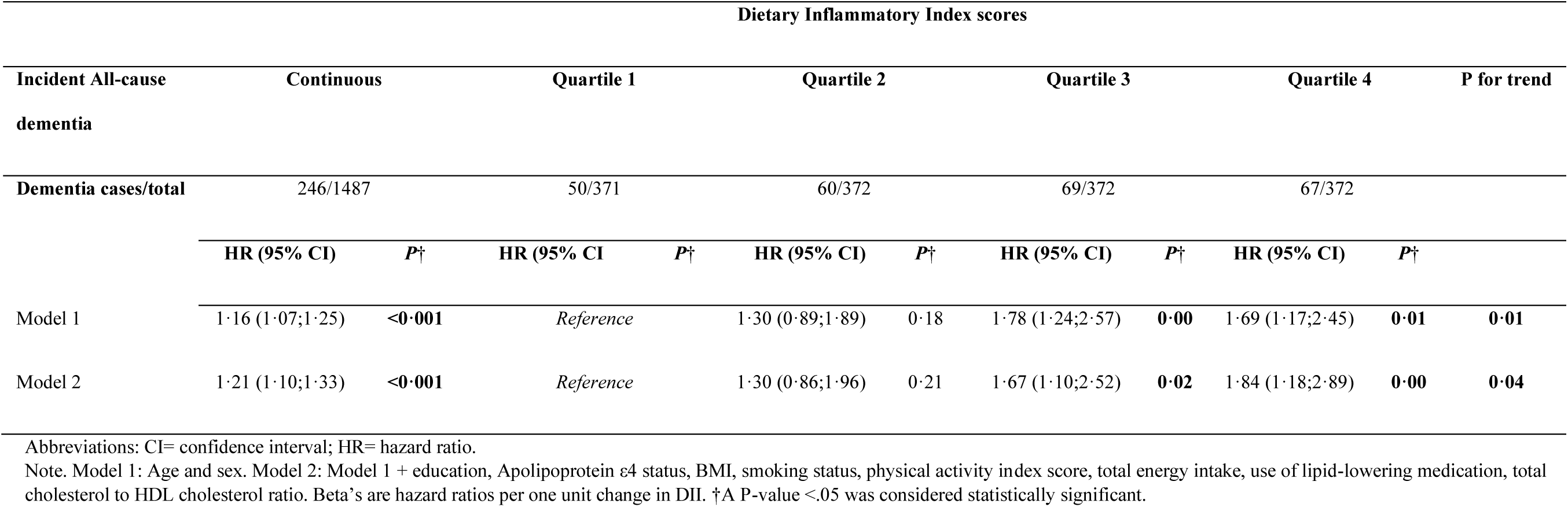
Dietary Inflammatory Index and Incident All-cause dementia (n=1487)

Similarly, higher DII scores were linearly associated with incident AD dementia (Model 1: 1·16 (1·06; 1·26), p= .001; Model 2: 1·20 (1·07; 1·34), p= .002) **(Table 3)**. In addition, in comparison to the first quartile, the third and fourth DII score quartiles were related with incident AD dementia in model 1 (Model 1: quartile 3: 1·71 (1·12; 2·62), p= .01; quartile 4: 1·74 (1·15; 2·65), p= .01), the relationship remained between the fourth DII score quartile, in comparison to the first quartile, and incident AD dementia after full adjustment (model 2: quartile 4: 1·89 (1·13; 3·17), p= .02).

**Table 3.** Dietary Inflammatory Index and Incident Alzheimer’s disease dementia (n=1487)

| Dietary Inflammatory Index scores |  |  |  |  |  |  |  |  |  |  |  |
| --- | --- | --- | --- | --- | --- | --- | --- | --- | --- | --- | --- |
| Incident Alzheimer's disease dementia | Continuous |  | Quartile 1 |  | Quartile 2 |  | Quartile 3 |  | Quartile 4 |  | P for trend |
| Dementia cases/total | 187/1487 |  | 38/ 371 |  | 47/372 |  | 50/372 |  | 52/372 |  |  |
|  | HR (95% CI) | P† | HR (95% CI) | P† | HR (95% CI) | P† | HR (95% CI) | P† | HR (95% CI) | P† |  |
| Model 1 | 1·16 (1·06;1·26) | **0·00** | Reference |  | 1·35 (0·88;2·06) | 0·17 | 1·71 (1·12;2·62) | **0·01** | 1·74 (1·15;2·65) | **0·01** | **0·04** |
| Model 2 | 1·20 (1·07;1·34) | **0·00** | Reference |  | 1·34 (0·84;2·14) | 0·23 | 1·50 (0·93;2·42) | 0·10 | 1·88 (1·13;3·15) | **0·02** | 0·11 |

Abbreviations: CI= confidence interval; HR= hazard ratio.
Note. Model 1: Age and sex. Model 2: Model 1 + education, Apolipoprotein ε4 status, BMI, smoking status, physical activity index score, total energy intake, use of lipid-lowering medication, total cholesterol to HDL cholesterol ratio. Beta's are hazard ratios per one unit change in DII. †A P-value <.05 was considered statistically significant.

We observed no significant interactions between DII scores and *APOE* ε4, sex, hypertension, diabetes mellitus type 2, and cardiovascular disease in association with incident all-cause and AD dementia **(eTable 1 in the Supplement)**.

## Discussion

In the present study we examined the relationships between DII scores and incidences of all-cause and AD dementia in the large community-based FHS Offspring cohort over a maximum of 22.3 years of follow-up. We found associations between higher pro-inflammatory DII scores and increased incidences of all-cause dementia and AD dementia. Our results may have important implications for diet related public health intervention strategies for dementia.

### Higher DII scores and incident all-cause and AD dementia

Our findings that higher DII scores were related with increased incidence of all-cause and AD dementia are in line with two previous studies.^16,23^ The Women’s Health Initiative Memory Study (WHIMS) reported that higher DII scores were associated with increased risk for probable all-cause dementia among women over 20 years of follow-up.^16^ Additionally, the Hellenic Longitudinal Investigation of Aging and Diet study (HELIAD) reported that higher DII scores were associated with an increased incidence of all-cause dementia over a maximum of 6.08 years of follow-up.^23^ Our investigation corroborated these findings, in addition we also examined the association with AD dementia, the most common form of dementia, confirming that the DII may be a risk factor contributing to Alzheimer’s disease pathology. Similar longitudinal studies examining this association in diverse populations across the world with unique dietary patterns would validate these findings.

### Mechanisms

Given the association that we detected, what is the likely pathophysiologic basis? One of the hypothesized contributary pathways leading to dementia is systemic inflammation.^10,11^ A driving factor of systemic inflammation is ‘inflammaging’, which activates microglial macrophages to produce pro-inflammatory cytokines (e.g. C-reactive protein, interleukin-1β, interleukin-6, and tumor necrosis factor-α).^11^ Production of these cytokines can cause neuronal death, cerebral small vessel disease, neurodegeneration, and reduce brain volume.^11^ In addition, another mechanism; beta-amyloid (Aß) induced injury in the brain, also stimulates the production of cytokines.^33^ In the brain cytokines contribute to an increase in Aß plaque formation, the hallmark of AD.^33^

### Diet, inflammation, and dementia implications

Multiple studies have shown that pro-inflammatory content in the diet is related to chronic systemic inflammation.^11^ The most pro-inflammatory components of the DII (i.e. saturated fat, trans fats, and total energy intake) are present in the pro-inflammatory “Western diet” - a summary term for dietary patterns consumed in western societies – which has shown association with increased concentrations of biomarkers of systemic inflammation and risk factors of all cause and AD dementia^34^, and also with neurodegenerative disease related outcomes.^34^ Western diets having inflammatory potential especially known to be pro-inflammatory may have enormous implications to preventive dietary interventions.

On the other hand, a diet which has shown to have potential to reduce systemic inflammation, to be beneficial for cardiometabolic health and promising to reduce all-cause dementia risk is the Mediterranean diet.^12,35^ Important findings from a study carried out by the Chicago Health and Aging Project study reported a significant relationship of higher Mediterranean diet score on slower cognitive decline only among participants with a low Western diet score and not among participants with a high Western diet score.^36^ This finding implies that the beneficial effect of the Mediterranean diet on health outcomes may depend on a low Western type dietary pattern score. Examining Western and Mediterranean dietary patterns alone may be limiting given the wide variety in dietary patterns consumed across the world. The DII is a summary measure that can capture inflammatory potential of the Western and Mediterranean and other dietary patterns, and may help generate dietary guidelines by public health policy makers.

RCTs have already shown associations between anti-inflammatory dietary patterns and neurodegenerative disease outcomes. Current clinical trial evidence is based on the PREDIMED trial carried out in Spain and a MIND diet RCT carried out in Iran.^7–9^ Both RCTs found beneficial effects of these dietary patterns on cognitive function. ^7–9^. New insights of an important RCT, which investigated the effect of the MIND diet on cognitive function and brain aging (n=604), are expected to be released soon (ClinicalTrials.gov NCT02817074).^37^ Trials have shown effects, which is encouraging. However, to have a sound understanding of how dietary interventions may help, elucidating the specific factors that influence the dementia process is important. Scoring the DII and other risk and protective factors in a given dietary intervention will help elucidate what specific features of the diet under study is influencing the pathology of the disease.

### Strengths and limitations

Strengths of this study include our large population-based sample, a maximum follow-up time of 22.3 years, a validated FFQ, and the ability to average dietary intake over a maximum of three time points to estimate dietary intake over a maximum of 10 years follow-up. In addition, we were able to adjust for many important confounders, including lifestyle and risk factors for all-cause and AD dementia.

However, we acknowledge that the present study has limitations. First, to assess dietary intake of our participants we used a FFQ, which is subject to measurement error and recall bias. To mitigate the effects of recall bias, we adjusted for total energy intake in model 2 to account for potential systematic measurement error.^38^ We acknowledge the possible presence of non-differential misclassification while using this dietary assessment method, which may have led to bias towards the null.^39^ Second, in the Framingham Heart Study Offspring cohort FFQ 36 out of 45 DII components were available. However, our quantity of components is in line with the two previous studies that investigated the DII and incident all-cause dementia (i.e. Hayden et al. 32 components; Charasis et al. 36 components).^16,23^ Third, the present study is observational, which precludes conclusions about the causality of the observed associations due to potential reverse causality. However, the DII score was constructed over three consecutive visits over a maximum of 10 years of follow up, which makes such a reverse causation bias less likely. Lastly, generalizability of our findings to other races/ethnicities may be limited as individuals included in our study were mainly white individuals of European ancestry. However, the DII accounts for that to a certain extent by including a mean and standard deviation of a representative world database, which we used in our calculation.^15^

In conclusion, in our community-based sample, higher DII scores were associated with increased incident all-cause and AD dementia. These findings are in line with previous studies which investigated the inflammatory potential of diet in relation to incident all-cause and AD dementia. We encourage future studies to investigate these relationships among diverse populations.

## Statements

### Data access

All authors confirm to have full access to all the data in the study and accept responsibility to submit for publication.

### Contributors

DMVL, HGM, MEJ, and JJH, conceptualization, wrote the statistical analysis plan, analyzed and interpreted the data, wrote the original draft of the manuscript, and revised the manuscript. DMVL and SS sourced the funding. AB and SS overlooked the study conduct, interpreted the data, and reviewed the manuscript for important intellectual content. MMG, HJA, JS, CY, MIS and PFJ reviewed the manuscript for important intellectual content. JJH and AB had access to the raw data. MEJ and JJH had final responsibility for the decision to submit for publication.

### Datasharing

Data are available on request for bone fide investigators from managing institution of the Framingham Heart Study https://www.framinghamheartstudy.org/fhs-for-researchers/.

## Acknowledgement

We thank the FHS participants for donating their time to our research.

