## Supplementary material for "Association between dietary inflammatory index score and incident dementia: results from the Framingham heart Study offspring cohort": Online Supplemental Material

**Supplementary eTable 1. Interaction in the association between DII score and incident All-cause dementia and Alzheimer's disease dementia (results are *P*-values)**

| | DII Score | | | | |
|---|---|---|---|---|---|
| | Interaction Variables | | | | |
| | **APOE ε4** | **Sex** | **JNC VII Stage I Hypertension** | **DM2** | **CVD** |
| Incident All-cause dementia | 0·96 | 0·73 | 0·45 | 0·54 | 0·55 |
| Incident Alzheimer's dementia | 0·50 | 0·96 | 0·34 | 0·97 | 0·58 |

Abbreviations: APOE ε4= Apolipoprotein ε4 status; HTN= The Seventh Report of the Joint National Committee on Prevention, Detection, Evaluation, and Treatment of High Blood Pressure Stage I hypertension; DM2= Diabetes Mellitus type 2; CVD= cardiovascular disease; DII= Dietary Inflammatory Index.
Note. Model 1: adjusted for age and sex.

**Supplementary eTable 2. Description of Dietary Inflammatory Index calculation**

| Description |
| --- |
| The DII is based on a set of literature-based dietary-component- specific inflammatory effect scores, dietary intake data from the population under study, and a representative world database that provides a mean and standard deviation for consumption of each dietary component in the global population.[1] These then become the multipliers to express an individual's exposure relative to the 'standard global mean' as a Z-score: subtracting the 'standard global mean' (i.e. from the world database) from the amount reported by the study participants and dividing this value by the world standard deviation. To minimize the effect of 'right skewing', this value is converted to a percentile score. To achieve a symmetrical distribution with values centered on 0 (null) and bounded between −1 (maximally anti- inflammatory) and +1 (maximally pro-inflammatory), each percentile score is doubled and then '1' is subtracted. The centered percentile value for each dietary component is then multiplied by its respective 'overall dietary component-specific inflammatory effect score' to obtain the 'dietary component specific DII score'. Finally, all of the 'dietary component-specific DII scores' are summed to create the 'overall DII score' for an individual, higher scores indicating pro-inflammatory DII scores).[1] |

NOTE. References


1. Shivappa N, Steck SE, Hurley TG, Hussey JR, Hebert JR. Designing and developing a literature-derived, population-based dietary inflammatory index. *Public Health Nutr* 2014; **17**(8): 1689-96.

**Supplementary eTable 3. Description of covariates**

| Covariate | Description |
|---|---|
| Apolipoprotein E ε4 allele | Apolipoprotein E ε4 allele genotyping was determined as described by Elosua et al. and Hixson and Vernier [1,2] |
| Body mass index | BMI was defined as weight (in kilograms) divided by the square of height (in meters). |
| Cholesterol | Fasting blood samples at examination cycle 7 were drawn from participants after an overnight fast. Total cholesterol and high-density lipoprotein (HDL) cholesterol concentrations were measured directly using standardized assays. |
| Education | Education was categorized into 3 groups (up to completion of 12 years of education leading to high school degree but no college; some college; college degree). |
| Lipid lowering medication | Lipid lowering medication (i.e. statins, fibrates, resins, niacin or nicotinic acid, and other anti-cholesterol drugs) usage was categorized into users and non-users. |
| Physical activity | Physical activity was self-reported using the physical activity index [3]. |
| Smoking status | Indicator variables were used for current smoking. |
| Total energy intake | Total energy intake was estimated from the Food Frequency questionnaire. |
| Prevalent cardiovascular disease | I.e. coronary heart disease, intermittent claudication, congestive heart failure, stroke, or transient ischemic attack. |
| Prevalent Diabetes Mellitus type 2 | Diabetes was defined as random blood glucose ≥200 mg/dL or fasting blood glucose ≥126 mg/dL or on anti-diabetic medication. |
| Prevalent hypertension | The Seventh Report of the Joint National Committee on Prevention, Detection, Evaluation, and Treatment of High Blood Pressure Stage 1 prevalent hypertension (i.e. systolic blood pressure ≥140mm Hg or diastolic blood pressure ≥90 mm Hg or current use of antihypertensive medications). |

NOTE. References


1. Elosua R, Ordovas JM, Cupples LA, et al. Association of APOE genotype with carotid atherosclerosis in men and women: the Framingham Heart Study. *J Lipid Res* 2004; **45**(10): 1868-75.
2. Hixson JE, Vernier DT. Restriction isotyping of human apolipoprotein E by gene amplification and cleavage with HhaI. *J Lipid Res* 1990; **31**(3): 545-8.
3. Kannel WB, Sorlie P. Some health benefits of physical activity. The Framingham Study. *Arch Intern Med* 1979; **139**(8): 857-61.
